# Kufor-Rakeb Syndrome-Associated Psychosis: A Novel Loss-of-Function *ATP13A2* Variant and Response to Treatment

**DOI:** 10.1101/2024.01.12.23300401

**Authors:** Mark Colijn, Stephanie Vrijsen, Ping Yee Billie Au, Rania Abu El Asrar, Marine Houdou, Chris Van den haute, Justyna Sarna, Greg Montgomery, Peter Vangheluwe

## Abstract

Biallelic (autosomal recessive) pathogenic variants in ATP13A2 cause a form of juvenile-onset parkinsonism, termed Kufor-Rakeb syndrome. In addition to motor symptoms, a variety of other neurological and psychiatric symptoms may occur in affected individuals, including supranuclear gaze palsy, spasticity, and cognitive decline. Although psychotic symptoms are often reported, response to antipsychotic therapy is not well described in previous case reports/series. As such, we describe treatment response in an individual with Kufor-Rakeb syndrome-associated psychosis. His disease was caused by a homozygous novel loss-of-function ATP13A2 variant (NM_022089.4, c.1970_1975del) that was characterized in this study. Our patient exhibited a good response to quetiapine monotherapy, which he has so far tolerated well. We also reviewed the literature and summarized all previous descriptions of antipsychotic treatment response. Although its use has infrequently been described in Kufor-Rakeb syndrome, quetiapine is commonly used in other degenerative parkinsonian disorders, given its lower propensity to cause extrapyramidal symptoms. As such, quetiapine should be considered in the treatment of Kufor-Rakeb syndrome-associated psychosis, when antipsychotic therapy is deemed necessary.

## Introduction

Biallelic pathogenic variants in ATP13A2, also known as PARK9, are associated with an autosomal recessive form of juvenile-onset parkinsonism, termed Kufor-Rakeb syndrome (KRS). Additional clinical features include facial-faucial-finger myoclonus, supranuclear gaze palsy, oculogyric dystonic spasms, and dementia, in addition to various neuropsychiatric symptoms, including psychosis [1, 2]. Some affected individuals are considered to have a form of neurodegeneration with brain iron accumulation or neuronal ceroid lipofuscinosis [1, 2], and spastic paraplegia has also been described in relation to recessive ATP13A2 variants [3].

*ATP13A2* encodes for the ATPase 13A2 (ATP13A2). ATP13A2 belongs to the superfamily of P-type ATPase transporters, which are alternatingly auto-phosphorylated and dephosphorylated on a conserved aspartic acid during their catalytic cycle [4]. ATP13A2 has recently been described as a polyamine transporter with highest affinity for the polyamines spermine and spermidine. At the cellular level, ATP13A2 is localized in late endolysosomes, where it exports the endocytosed polyamines spermine and spermidine from the lumen to the cytosol [5]. Polyamines are neuroprotective agents and are involved in a plethora of pathways, ranging from the regulation of transcription and translation to autophagy and anti-oxidant responses [6]. As ion channel modulators, polyamines have been implicated in several mental disorders, including schizophrenia, mood disorders, anxiety, and suicidal behaviour [7]. ATP13A2 loss-of-function causes a disturbed intracellular polyamine distribution with (i) polyamine accumulation in late endolysosomes, resulting in rupture of these organelles and cathepsin B-mediated cell death [5], and (ii) a deficiency of cytosolic polyamines, leading to more mitochondrial derived reactive oxygen species causing oxidative stress [8]. Interestingly, all KRS variants characterized up until now show a (nearly) complete loss of ATP13A2 function as a consequence of protein mislocalization, instability, hampered autophosphorylation, dephosphorylation, and/or ATPase activity, or a combination thereof [9–13, 3, 5].

Although psychotic symptoms may occur in individuals with KRS (even in the absence of dopaminergic therapy), response to antipsychotic treatment has infrequently been described in the literature. As particular recommendations regarding the treatment of psychosis in this population remain scarce, we report a proband with KRS-associated psychosis caused by a homozygous novel loss-of-function *ATP13A2* variant, who ultimately responded well to quetiapine monotherapy. We also performed a review of the literature with respect to treatment response in individuals with psychosis.

### Case Report

We report a young adult male with a mild intellectual disability (FSIQ=73 in 2016) who presented to clinical attention after abruptly developing psychotic symptoms in his teenage years. He was born around 36 weeks gestation via cesarean section due to either oligohydramnios or breech position (there have been conflicting reports in this respect). Although there were no gross motor delays he was described as being clumsier than his peers and later struggled with handwriting. He was delayed in both expressive and receptive speech and had articulation difficulties. Although he did not undergo a formal psychoeducational assessment until his teenage years, he reportedly always struggled academically. Starting at around the time of his psychiatric admission (but possibly earlier), his cognitive abilities began to progressively deteriorate. He nonetheless completed grade 12. Frank motor symptoms had not been identified or endorsed prior to the initiation of antipsychotic medication (see below for details).

At the time of his initial assessment in a specialized movement disorders clinic in the years following his admission to hospital, he exhibited new-onset marked difficulties with vertical eye movements in the upward plane with hypometric saccades in the horizontal plane. There was no blepharospasm and no orofacial dystonia. Marked hypomimia and hypophonia were noted. Moderate rigidity was noted axially as well as in both upper extremities with marked (slightly asymmetric) bradykinesia in all four limbs (more prominent in the left upper extremity and right lower extremity). He had no difficulty standing up from a seated position. Gait examination revealed markedly reduced stride length and arm swing, particularly in the left upper extremity. The pull test was positive. Deep tendon reflexes were brisk in the upper extremities at 3+ with no Hoffmann’s sign, and 2+ at the knees. No spasticity was appreciated. No tremor or any other abnormal movements were observed.

Apart from vaguely described anxiety, of a generalized and social nature, as well as possible attention-deficit/hyperactivity disorder, he was generally psychiatrically well prior to his index admission to hospital, which occurred immediately following a traumatic event. Although his symptoms were initially queried to represent a severe trauma reaction, the presence of frank psychotic symptoms eventually became evident, including disorganization of thought and behaviour, as well as persecutory, referential, and hyperreligious delusions that occurred in association with thematically congruent auditory hallucinations. While his psychotic symptoms initially resolved with the initiation of low dose risperidone, he experienced remarkably severe extrapyramidal symptoms (EPS). As such, he was transitioned to olanzapine, which was comparatively less problematic from a motor side effect standpoint, but nonetheless continued to cause atypically severe EPS on doses as low as 2.5 mg qhs.

His overall neuropsychiatric and developmental history prompted a medical genetics service consult. While initial testing with chromosomal microarray was negative, clinical exome sequencing (Blueprint Genetics 2021) revealed a homozygous variant in ATP13A2 (NM_022089.4, c.1970_1975del) which is predicted to cause an in-frame deletion of two amino acids, p.(Pro657_Glu658del). This variant is rare and absent from the gnomAD v4 database (accessed Nov 28, 2023). Although this in-frame deletion was initially formally classified as a variant of uncertain significance, it was clinically considered to be likely pathogenic given that his overall presentation was highly suggestive of KRS. Additionally, biochemical analysis demonstrating that the variant causes loss-of-function was performed (see below for details), confirming pathogenicity. His workup was otherwise unrevealing, including a brain MRI which did not show obvious evidence of neuronal brain iron accumulation.

While his psychotic symptoms and comorbid anxiety were initially reasonably well controlled with low dose olanzapine following his discharge from hospital, he exhibited worsening dysarthria, dysphagia, bradykinesia, and balance difficulties over the subsequent years. When his olanzapine dose was decreased to 1.25 mg qhs, his motor symptoms improved but he became more paranoid and anxious. As such, he was eventually cross-titrated from olanzapine to quetiapine. Although he experienced a reemergence of symptoms at certain points during the transition, he eventually again achieved remission of his psychotic symptoms on quetiapine monotherapy (quetiapine XR 200 mg qhs in addition to quetiapine IR 100 mg qhs). A combination of the extended release and immediate release formulations was chosen, as he initially experienced breakthrough symptoms in a diurnal pattern (specifically, in the evenings prior to his next dose) on the immediate release formulation alone. Additionally, as expected his motor symptoms noticeably improved with this change, despite some degree of ongoing dysarthria, rigidity, and bradykinesia, as well as continued reduced stride length and arm swing while walking. On recent examinations facial-faucial-finger mini-myoclonus has also been observed.

## Methods

### Biochemical Analysis of the ATP13A2 Variant

#### Cell culture

Human neuroblastoma SH-SY5Y cell lines either non-transduced (nts) or stably overexpressing wild-type ATP13A2, a catalytically dead variant (D508N), or the Pro652_Glu653del ATP13A2 variant (the nomenclature is based on the sequence of ATP13A2 splice variant 2, which is 5 amino acids shorter at the N-terminal region and has been historically used for biochemical analysis [5]) were generated via lentiviral transduction as described previously [14]. A detailed protocol can be found at dx.doi.org/10.17504/protocols.io.bw57pg9n. Addgene ID’s used were: #213697 (WT), #171820 (D508N) and #213700 (Pro652_Glu653del). Cells were maintained at 37 °C in the presence of 5% CO2 and incubated in high-glucose Dulbecco’s modified Eagle medium supplemented with 1% penicillin/streptomycin (Sigma), 15% fetal calf serum (heat inactivated) (Sigma), 1% non-essential amino acids (Sigma), 1% sodium pyruvate (Gibco), and selection antibiotic (160 µg/mL hygromycin or 2 µg/mL puromycin (Invivogen)). The treatments were performed in the same medium, with the exclusion of selection antibiotic.

#### Immunofluorescence

Cells were seeded at 75 000 cells/well in a 12-well plate with coverslips and allowed to attach and grow for 48 h. Thereafter, cells were washed with ice-cold PBS and fixed with 4% paraformaldehyde (Thermo Fisher Scientific) (30 min, 37 °C). Cells were washed twice more with ice-cold PBS before permeabilization and blocking with a mixture of 5% BSA (Roth) and 0.5% saponin (Sigma) (referred to as blocking buffer) for 1 h. Next, cells were incubated with primary antibody (anti-ATP13A2, A3361, Sigma; anti-CD63, 11-343-C100, ExBio) (diluted 1/200 in blocking buffer) for 2 h and then subjected to secondary antibody (Alexa-488 goat anti-rabbit IgG, R37116, Invitrogen; Alexa-594 goat anti-mouse IgG, A11005, Invitrogen) (diluted 1/1000 in blocking buffer) for 30 min on a shaker. Subsequently, cells were immersed in 200 ng/ml DAPI (D9542, Sigma) for 15 min. Samples were thoroughly washed with PBS in between the different steps. Finally, the samples were mounted and images were acquired using a LSM880 microscope (Zeiss) with a 63x objective and Airyscan detector.

#### SDS-page and immunoblotting

70% confluent cells were harvested and subsequently lysed with radio-immunoprecipitation assay buffer (Thermo Fisher Scientific) supplemented with protease inhibitors (Sigma). 10 µg of protein (concentration determined with a bicinchoninic acid protein assay) was then loaded on a NuPage 4-12% Bis-Tris gel (Thermo Fisher Scientific, Bio-Rad) and subjected to an electrophoresis run at 130 V in MES running buffer (Thermo Fisher Scientific). The proteins were subsequently transferred to a PVDF membrane in NuPage transfer buffer (Thermo Fisher Scientific) supplemented with 10% v/v methanol (Roth). Immunoblots were blocked by incubation in 5% w/v milk powder (1 h, room temperature). Next, immunoblots were probed with primary antibodies (anti-GAPDH, G8795, Sigma; anti-ATP13A2, A3361, Sigma) (diluted 1/5000 and 1/1000 in 1% w/v BSA, respectively) and incubated O/N (4 °C). After thorough washing, immunoblots were subjected to secondary antibodies (HRP-linked anti-mouse IgG, 7076S, Cell Signaling; HRP-linked anti-rabbit IgG, 7074S, Cell Signaling) (diluted 1/2000 in 1% w/v milk powder) (1 h, room temperature). All dilutions and wash steps were performed with TBS supplemented with 0.1% v/v Tween-20 (PanReac AppliChem). Detection was performed by means of chemiluminescence (Bio-Rad ChemiDoc) and protein levels were quantified with Image Lab software.

#### ^14^C-labeled polyamine uptake

This protocol is based on a previous publication [15], with minor modifications. Briefly, 70% confluent SH-SY5Y cells in a 12-well plate were incubated (30 min, 37 °C) either with 5 µM ^14^C-labeled spermine (3139-50 µCi, ARC) or with a mixture of 5 µM ^14^C-spermine and 100 µM unlabeled spermine in cell culture medium. The medium was subsequently aspirated and cells were washed twice with ice-cold PBS (without Ca^2+^ and Mg^2+^). Next, the cells were lysed by incubation in 200 µl radio-immunoprecipitation assay buffer (10 min,room temperature) (Thermo Fisher Scientific) before scraping. The cell lysate was added to scintillation vials filled with 7 ml EcoLite Liquid Scintillation Cocktail (01882475-CF,MP Biomedicals). Thereafter, the wells were washed with 200 µl ice-cold PBS (without Ca^2+^ and Mg^2+^), which was added to the accompanying scintillation vial. Finally, ^14^C radioactivity in counts-per-minute (CPM) was measured with liquid scintillation counting (TRI-CARB 4910TR V Liquid Scintillation Counter, PerkinElmer).

#### Microsome collection

SH-SY5Y cells overexpressing wild-type or mutant ATP13A2 were seeded in 500 cm^2^ plates. Once they reached 70-80% confluency, cells were collected. Next, cells were lysed by resuspending the cell pellet in 3 ml hypotonic LIS buffer (10 mM Tris.HCl pH 7.5, 0.5 mM MgCl_2_.6H_2_O, 1 mM DTT, 1x SigmaFast protease inhibitors) (S8830, Merck), which was incubated on ice for 15 min. The suspension was transferred to a Dounce homogenizer and 60 up-and-down strokes were applied, followed by addition of 3 ml 1 M solution (0.5 M sucrose, 10 mM Tris.HCl pH 7.3, 40 µM CaCl_2_, 1 mM DTT, 1x SigmaFast protease inhibitors) and another 30 up-and-down strokes. The nuclear (1000 x g, 10 min, 4 °C), mitochondrial-lysosomal (12 000 x g, 20 min, 4 °C), and microsomal fractions (140000 x g, 35 min, 4 °C) were then collected. Fractions were suspended in 0.25 M sucrose with 1x SigmaFast protease inhibitors.

### ADP-Glo assay

ATPase activity was assessed using a commercially available luminescence assay (ADP-Glo Max assay, V7002, Promega) according to manufacturer’s instructions. A reaction mixture (final volume 25 µl) was made in a 96-well plate and contained 50 mM MOPS-KOH (pH 7), 100 mM KCl, 11 mM MgCl_2_, 1 mM DTT, 0.1 mg/ml DDM, 5 µg microsomes (1:2 ratio DDM: microsomes) and various concentrations of polyamines. The microsomes were collected from SH-SY5Y cells overexpressing ATP13A2 (wild-type or mutants). Next, the reaction was incubated (20 min, 37IT°C followed by 10 min, room temperature) following the addition of 5 mM ATP and terminated by adding 25 μl of ADP-Glo Reagent. The 96-well plate was subsequently incubated (40 min, room temperature), followed by the addition of 50 μl of ADP-Glo Detection Reagent. After 60 min, luminescence was detected using the Bio Tek plate reader.

### RNA collection and qPCR

RNA was isolated using the NucleoSpin RNA plus kit (Macherey-Nagel) following manufacturer’s instructions. RNA concentration and purity were determined using a Nanodrop spectrometer (Thermo Fisher Scientific). 5 µg RNA was then converted to cDNA using the High-Capacity cDNA Reverse Transcription Kit (Thermo Fisher Scientific). A 96-well plate was prepared with a ten-fold dilution of each cDNA sample in duplicates (with a volume of 5 µl cDNA per well), to which the following reaction mixture was added (volumes are given per well): 10 µl SYBR Green master mix (Roche), 1 µl of 5 µM forward primer, 1 µl of 5 µM reverse primer, and 3 µl distilled water. The cDNA was replaced by an equivalent volume of distilled water for the negative controls. Primers targeting the ATP13A2 gene were 5’-CATGGCTCTGTACAGCCTGA-3 ‘(forward) and 5’ CTCATGAGCACTGCCACTGT-3 ‘(reverse). Primers targeting the β-actin gene were 5’-CACTGAGCGAGGCTACAGCTT-3 ‘(forward) and 5’-TTGATGTCGCGCACGATTT-3 ‘(reverse). The qPCR read-out was performed, in which the reaction was initiated at 95 °C for 10 min, followed by 50 cycles at 95 °C for 10 sec and 55 °C for 30 sec, and ended at 95 °C for 1 min. A melting curve was determined from 55 to 95 °C.

## Literature Search

PubMed and Embase searches were completed in April 2023 (and updated in December 2023) using the terms “kufor-rakeb”, “kufor rakeb”, “KRPPD”, “parkinson disease-9”, “parkinson disease 9”, “PARK9”, “pallido-pyramidal degeneration with supranuclear upgaze paresis and dementia”, “ATP13A2”, or “1p36.13”, in combination with “schizophrenia”, “psychosis”, “psychotic”, “hallucinations”, “delusions”, “paranoia”, “antipsychotic”, or “neuroleptic”. The Pubmed search yielded nine results and the Embase search yielded 26. After duplicates were removed, 33 results remained. All results were manually reviewed, including their reference lists, for additional relevant articles. OMIM was also reviewed. Only articles that described the use of antipsychotic medications in individuals with Kufor-Rakeb syndrome were included in the review. The search process is outlined in Figure 1.

**Figure 1.**
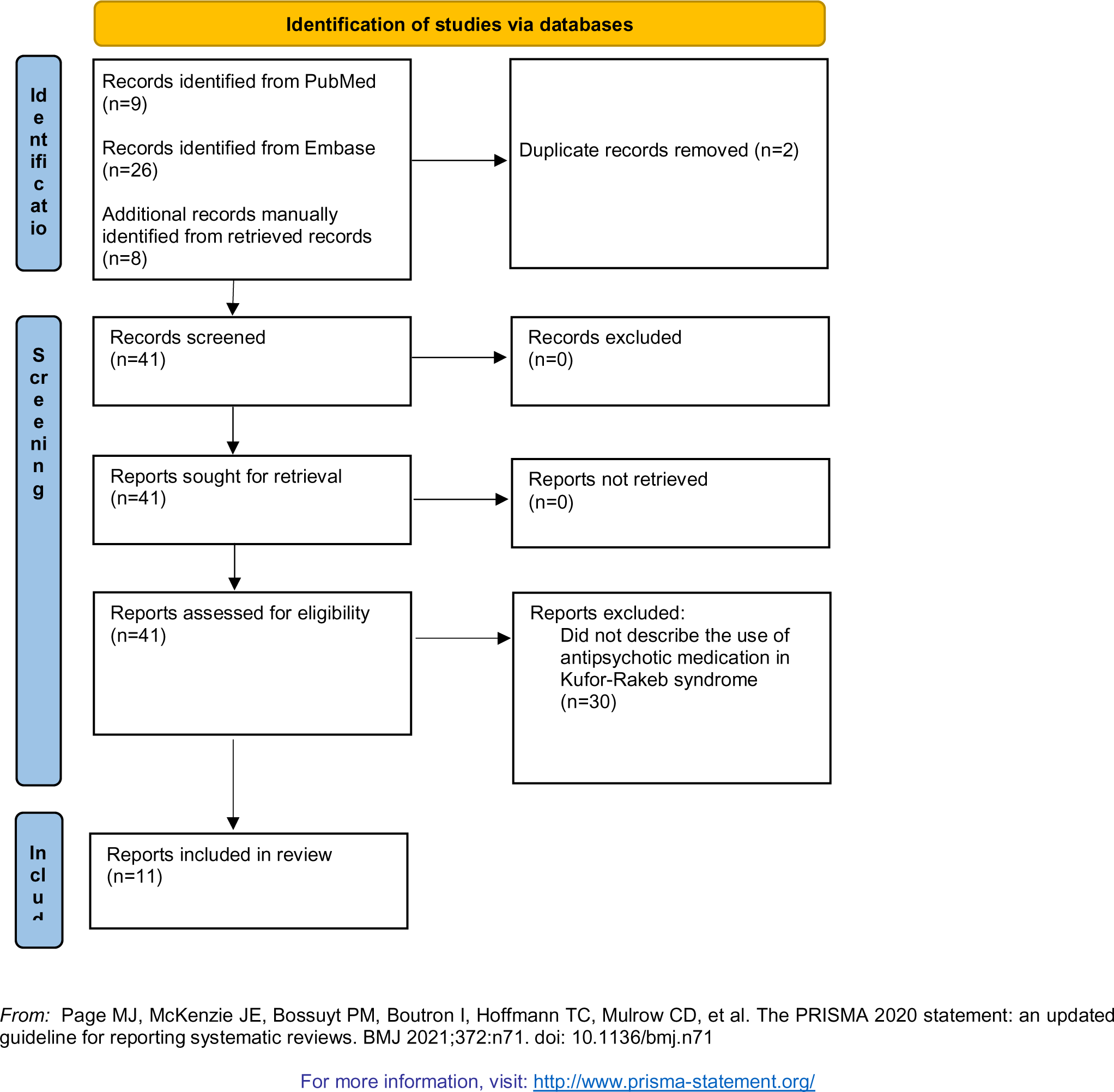
Literature Search Flow Diagram for Antipsychotic Use in Kufor-Rakeb Syndrome.

## Results

### Biochemical Analysis of the ATP13A2 Variant

To functionally characterize the novel ATP13A2 variant described in this study, we generated SH-SY5Y neuroblastoma cells overexpressing either wild-type ATP13A2, a catalytically dead variant that is defective in auto-phosphorylation (D508N, functioning as a negative control) or the Pro652_Glu653del variant. Cells overexpressing Pro652_Glu653del presented a significantly lower uptake of ^14^C-labeled spermine as compared to cells overexpressing wild-type ATP13A2 (Figure 2A), which offers a read-out for ATP13A2’s transport activity. This correlated well with the spermine-dependent ATPase activity in the membrane fractions of these cell lines. The ATPase activity of the Pro652_Glu653del protein was comparable to the D508N loss-of-function mutant pointing to complete loss of activity (Figure 2B). Whereas wild-type and D508N ATP13A2 colocalized with CD63, a late endolysosomal marker, the Pro652_Glu653del variant displayed a mesh-like pattern suggesting mislocalization in the endoplasmic reticulum (Figure 2C). We also analyzed the expression level, and found that Pro652_Glu653del mRNA levels were significantly higher than wild-type (Figure 2D), demonstrating successful transduction. In contrast, the Pro652_Glu653del variant exhibited a lower protein expression than wild-type or D508N ATP13A2 (Figure 2E), suggesting that protein stability may be impaired. In conclusion, in line with other previously described KRS variants [5], the Pro652_Glu653del variant exhibits full loss-of-function, which can be explained by a combination of protein instability, mislocalization, and inactivity. Our biochemical analysis indicates that in a homozygous context, this variant may be disease causing.

**Figure. 2.**
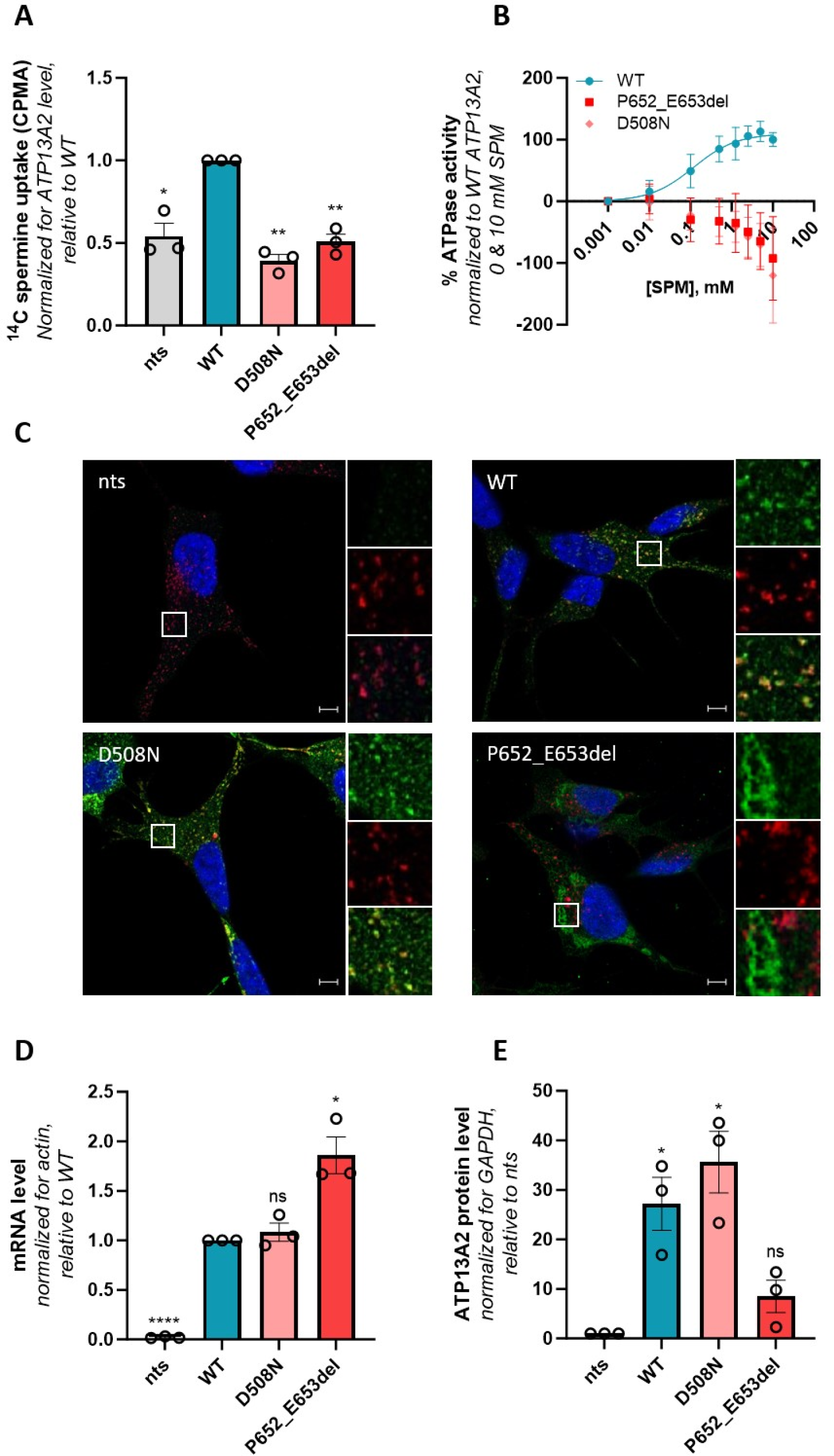
Functional characterization of the Pro652_Glu653del *ATP13A2* variant. SH-SY5Y neuroblastoma cells either non-transduced (nts) or stably transduced with constructs encoding for wild-type ATP13A2 (WT), a catalytically dead variant (D508N), or the Pro652_Glu653del (P652_E653del) ATP13A2 variant were used to analyze (A) cellular polyamine uptake potential by scintillation counting following a 30 min incubation wit^14^hC-labeled spermine. CPMA, counts-per-minute (n=3).(B) Spermine-induced ATPase activity was determined in isolated microsomes (n=3-4). Fitting was performed using the non-linear allosteric sigmoidal(.C) Colocalization of ATP13A2 (in green) with the late endolysosomal marker CD63 (in red) was analyzed by immunofluorescence (n=3). Scale bar, 5 µm. Individual and merged channels are shown for the boxed areas. ATP13A2 mRNA (n=3(D)) and protein (n=3)(E) levels in the SH-SY5Y cells were assessed via immunoblotting and qPCR, respectively. *, p<0.05; **, p<0.01; ns, non-significant versus WT (A, D) or nts (E); one-sample t-test.

### Literature Search

Although psychotic symptoms have often been described in KRS, only six articles (described below) have reported on the effectiveness of antipsychotic therapy in affected individuals. The developmental and neurological histories of these individuals, as well as information regarding their particular ATP13A2 variants, are summarized in Table 1. Five additional articles alluded to the use of antipsychotic medication but did not provide any information regarding treatment response [16–20]. The clinical details pertaining to the psychiatric aspects of these additional five cases are outlined in Table 2.

**Table 1.**
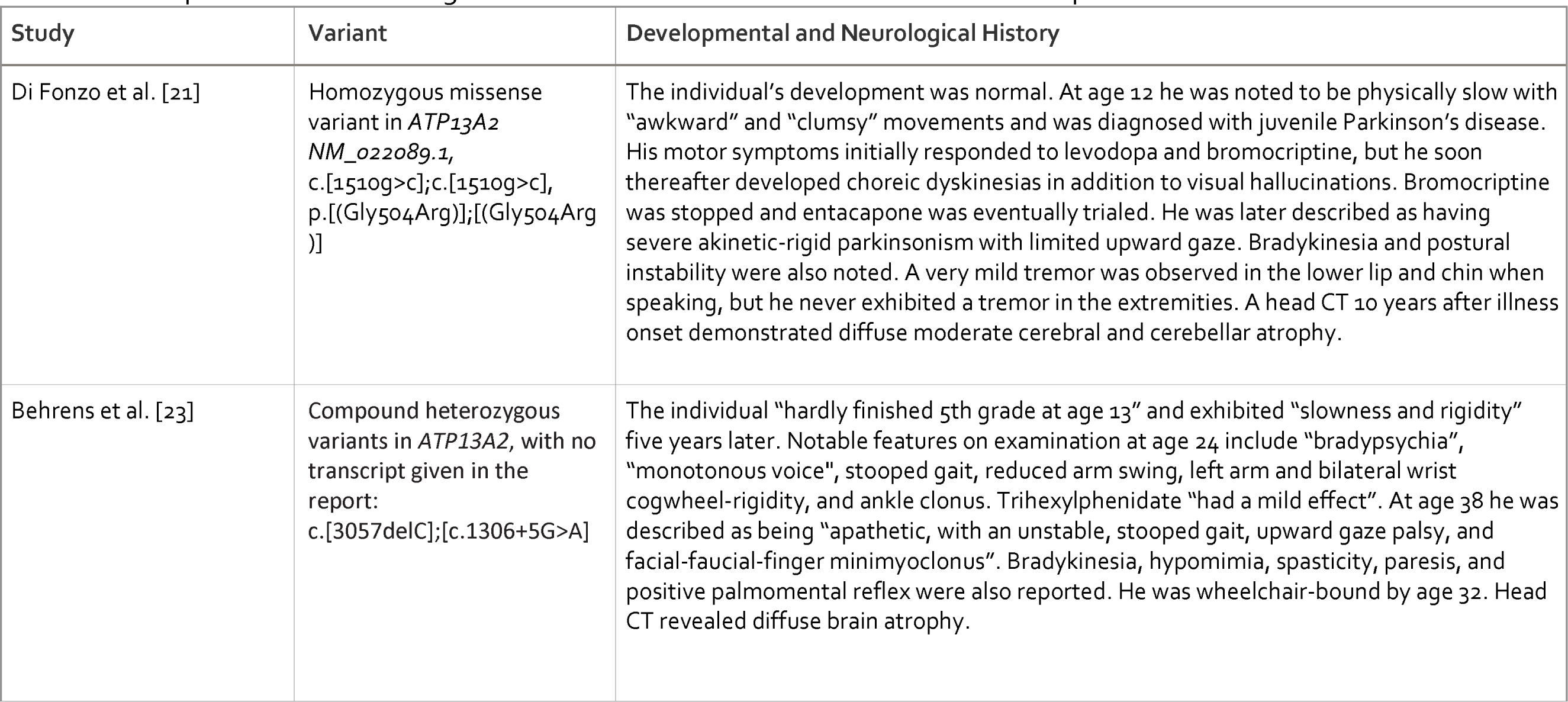
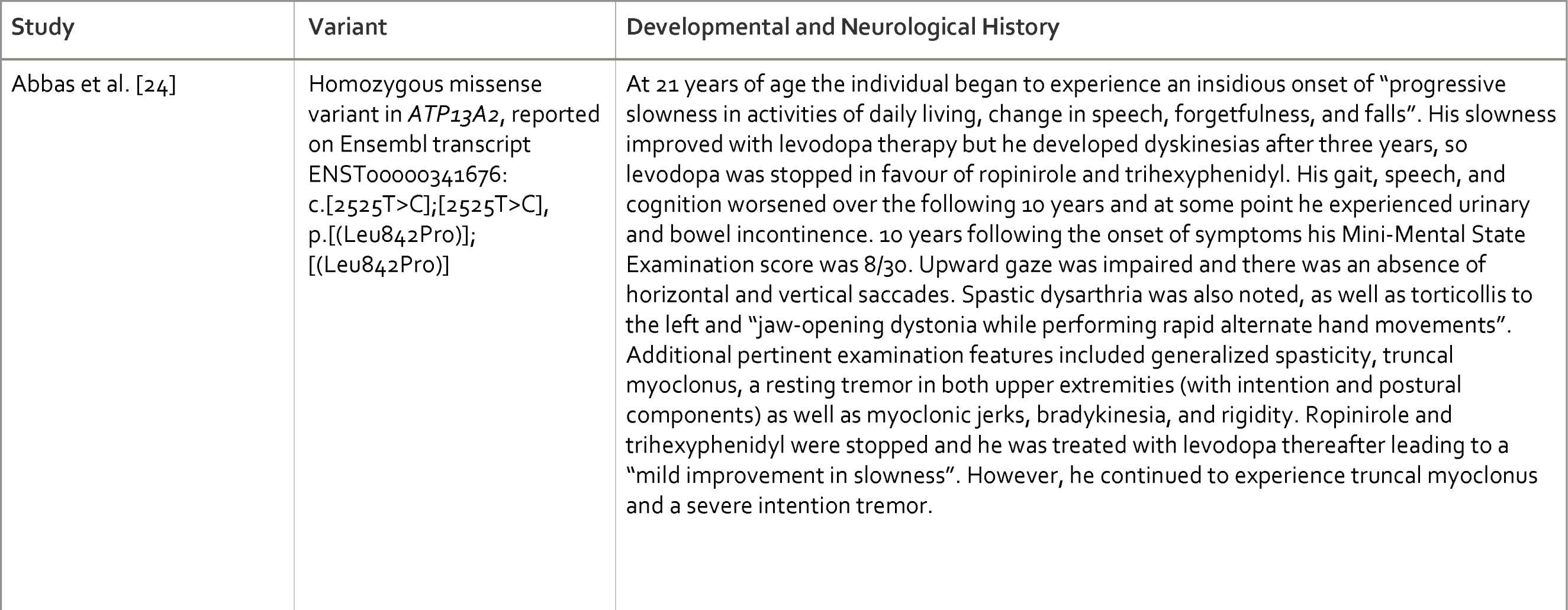
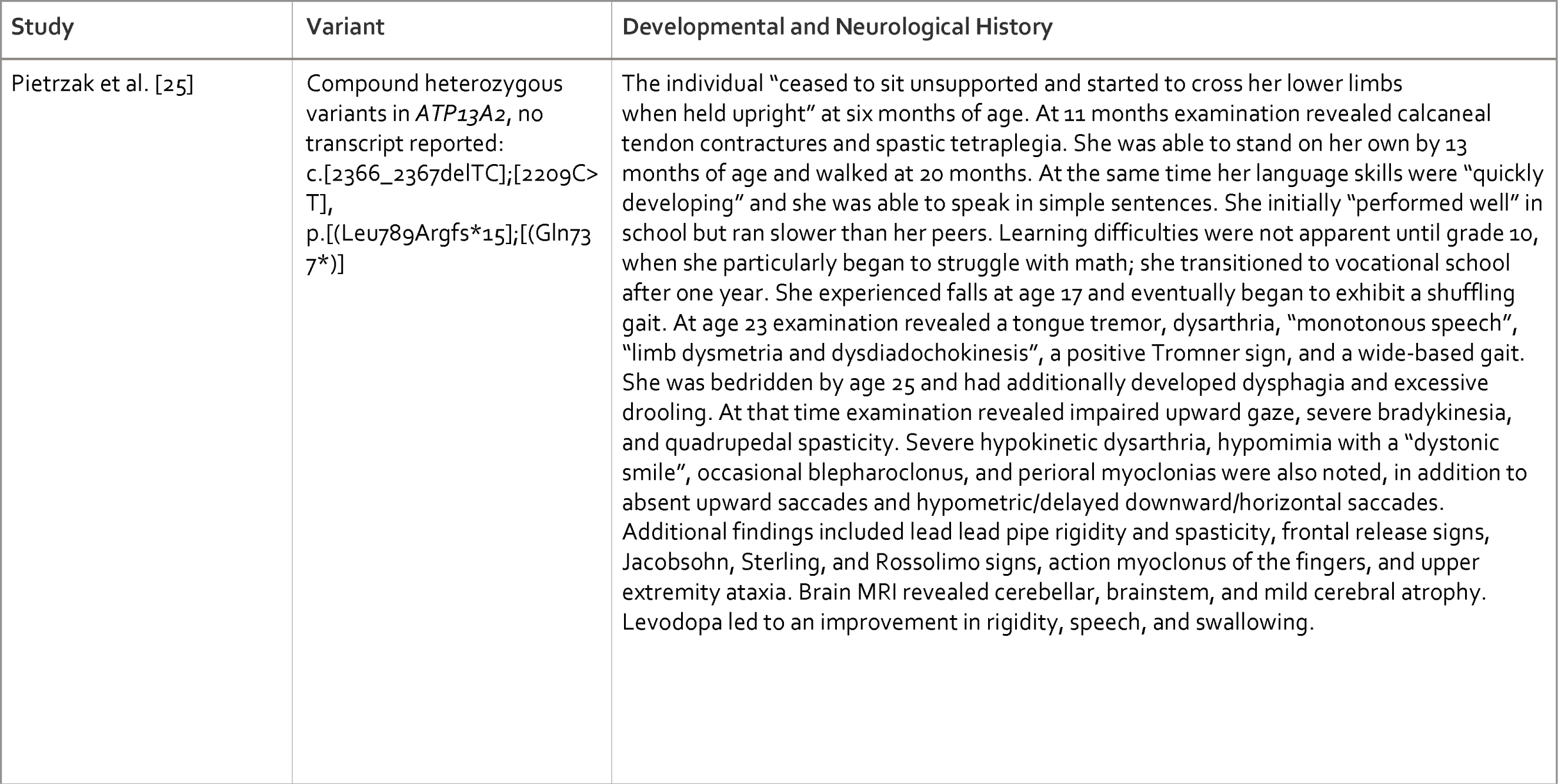
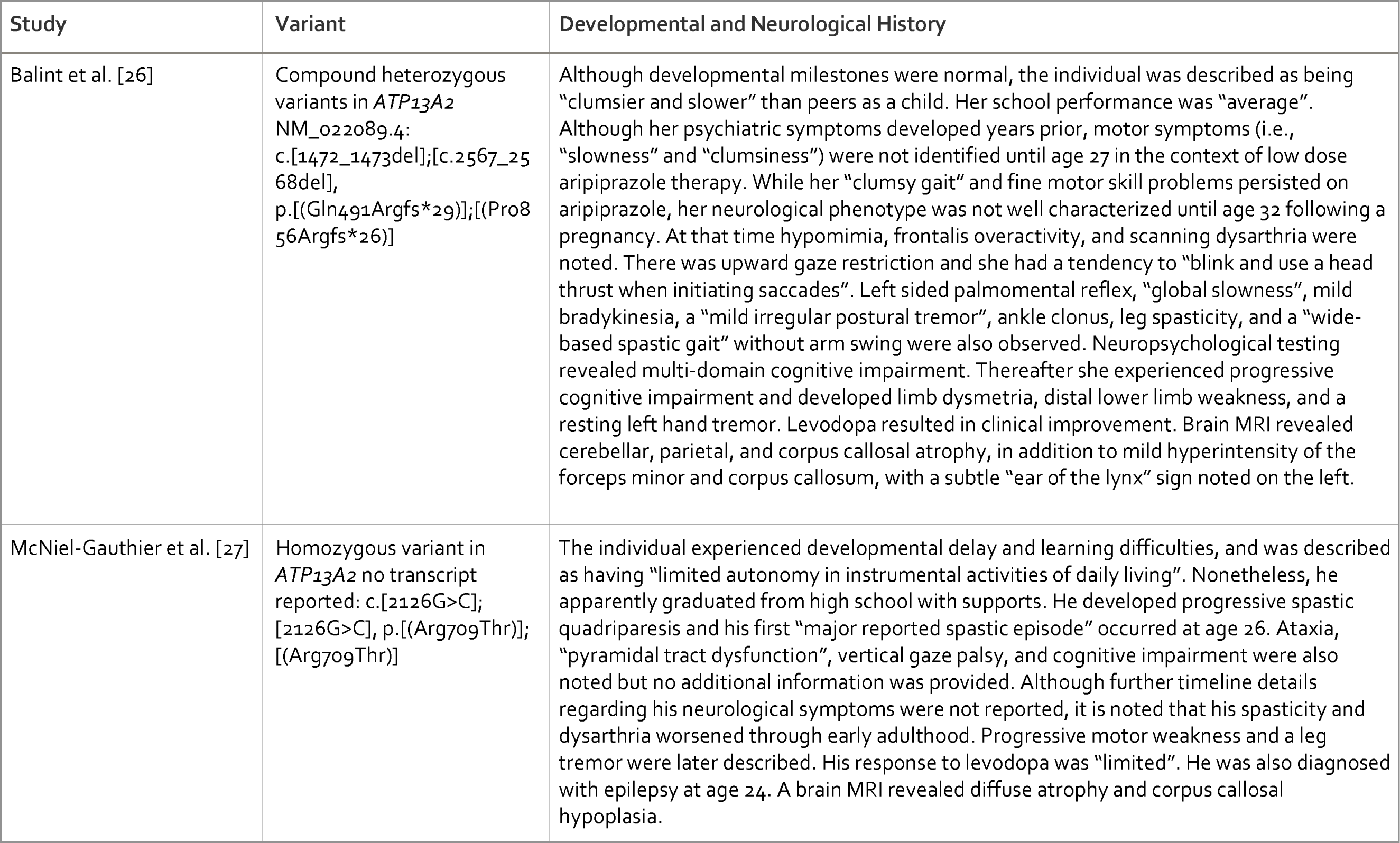
Developmental and Neurological Features in Cases that Describe Treatment Response.

**Table 2.**
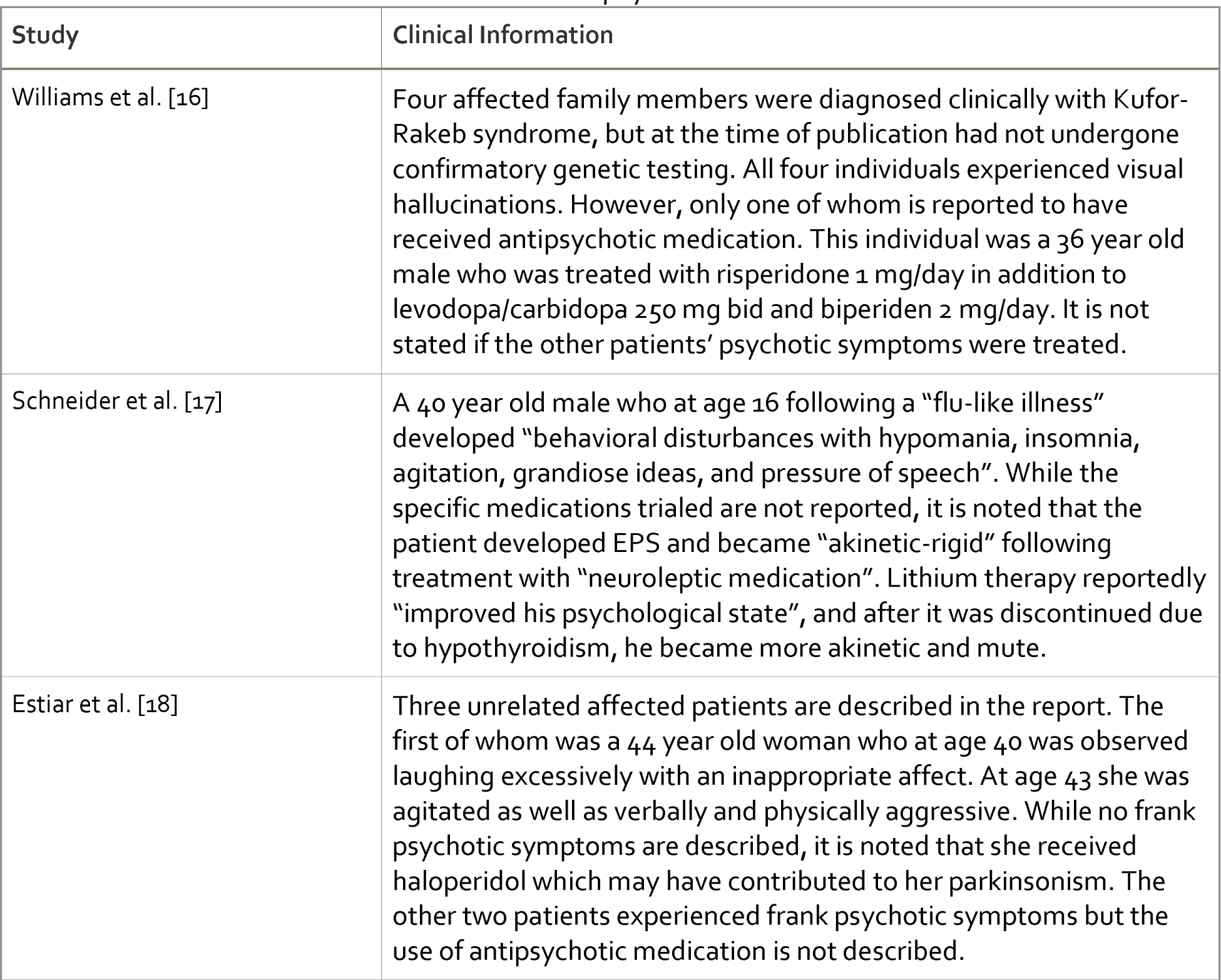
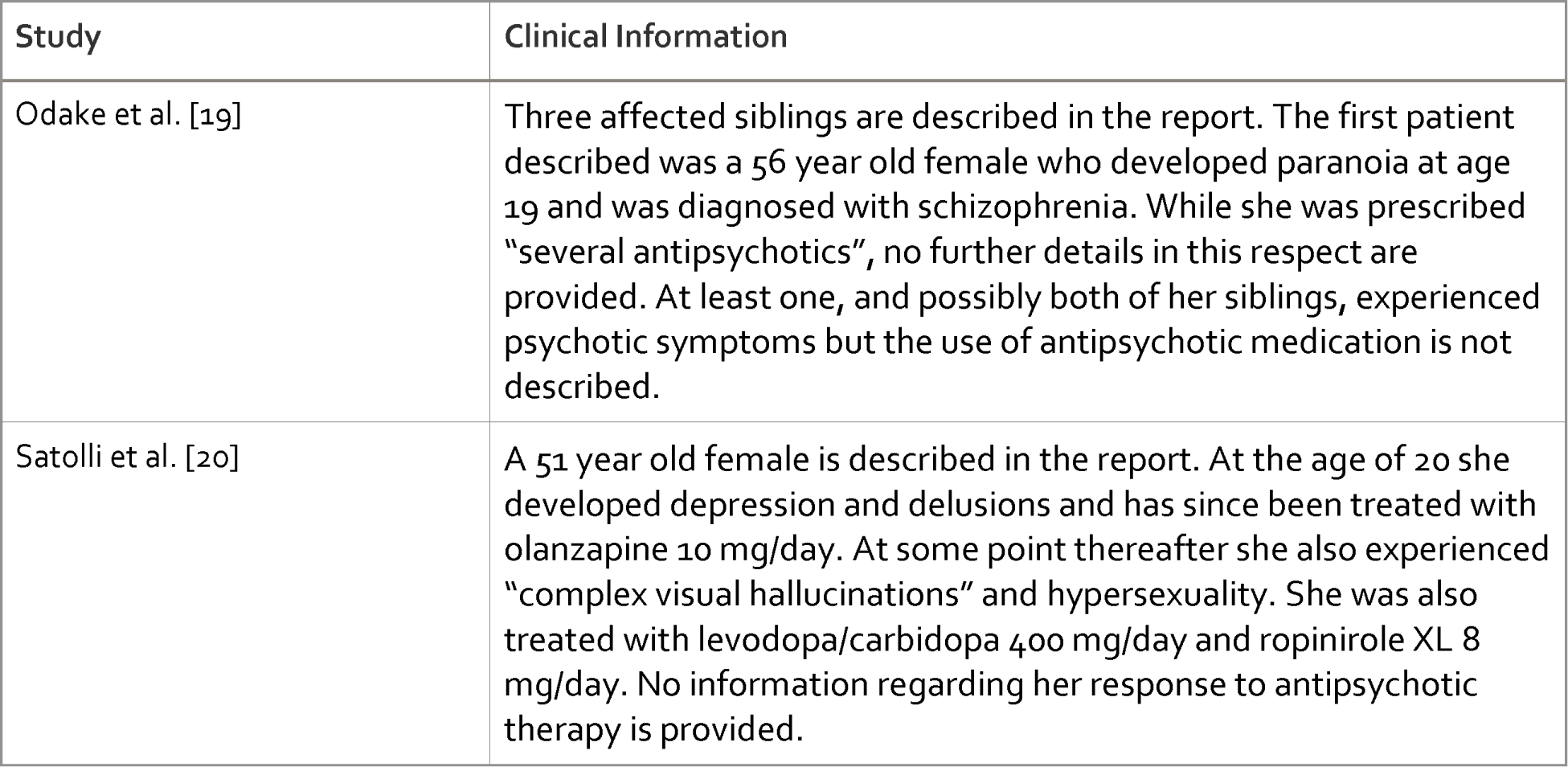
Additional Articles that Reference Antipsychotic Use.

Di Fonzo et al. [21] were the first to describe the treatment of psychotic symptoms in a patient with KRS; a male who developed visual hallucinations, and eventually aggression, as a young adult after the initiation of levodopa and bromocriptine. While he was admitted to hospital and treated with haloperidol, the effects it had on his psychotic and motor symptoms were not described. Nonetheless, it is stated that bromocriptine was stopped and a combination of levodopa/benserazide and quetiapine “allowed satisfactory control” of his motor and psychotic symptoms. However, during the year prior to his assessment by the authors of the report, his parkinsonism worsened “with re-emergence of hallucinations and severe fluctuations and dyskinesias”, requiring changes to his medication regimen. At the time of the assessment he continued to experience “psychiatric disturbances” during “on” states and his medications included levodopa (plus benserazide) 350 mg/day and quetiapine 50 mg/day. Following his death, a subsequent paper described this individual’s neuropathological characteristics, but no additional information regarding his psychiatric phenotype or response to treatment was provided [22].

Behrens et al. [23] described numerous members of an affected family, including one who developed episodes of confusion and “paranoid auditory hallucinations” in adulthood, seemingly in response to trihexylphenidate therapy. The occurrence of visual hallucinations is also mentioned in a table but not described in detail. Thioridazine 25 mg/day “improved” his hallucinations but worsened his EPS. While two siblings also experienced hallucinations, it is not reported if they were treated with antipsychotic medication.

Abbas et al. [24] described a 32 year old male who developed “excessive fear” as well as auditory and visual hallucinations at age 27, in addition to hypersexuality at age 29, in the context of longstanding ropinirole therapy. While it is noted that he had been treated with risperidone, olanzapine, quetiapine, and levosulpiride, his response to these medications is not described. At 32 years of age the initiation of clozapine 25 mg/day (in addition to clonazepam 2 mg/day) led to a reduction in his hallucinations, despite ongoing treatment with levodopa/carbidopa 200/50 mg tid (ropinirole had been stopped).

Pietrzak et al. [25] described a 28 year old female who at age 25 developed psychosis characterized by “persecutory delusions of a poorly defined imminent threat and auditory hallucinations (scary voices)”. Her symptoms were reportedly “controlled with olanzapine”. At the time of the initial assessment by the authors, her medication regimen included levodopa with benserazide “50 mg + 12.5 mg tid”, quetiapine 25 mg bid, and olanzapine 2.5 mg/day. While her levodopa was increased to “100 mg + 25 mg qid”, “an attempt at further increase” resulted in hypotension and a return of persecutory delusions. Paroxetine was also prescribed for anxiety.

Balint et al. [26] described a 38 year old female who developed paranoia at age 24, characterized by delusions of infidelity involving her husband as well as a belief that her work colleagues were “going through her notes” and talking “behind her back”. She was also diagnosed with bulimia nervosa at some point in adulthood. Her symptoms worsened at age 27, such that she “developed psychosis with complex persecutory delusions with episodes of visual hallucinations”. Notably, she was not receiving dopaminergic therapy at this time, as she had not yet been diagnosed with KRS. “Antidepressants and neuroleptics” caused “sleepiness, slowness, and clumsiness”, even at low doses. Although her “psychiatric symptoms” waxed and waned while her medication was adjusted, her response to treatment during this period is not described in detail. While aripiprazole eventually “stabilized her”, it is unclear if her psychotic symptoms fully remitted. Although her maximum dose of aripiprazole is not specified, at age 32 she was taking 2.5 mg/day. Aripiprazole was stopped shortly thereafter (following the diagnosis of KRS) which led to a mild improvement in her “slowness”. However, it is not reported if this change also led to an improvement in her other motor symptoms or a worsening of her psychotic symptoms. However, the authors do state that the initiation of levodopa/carbidopa 300 mg/day led to an improvement in her tremor and bradykinesia, and that from age 32 onward she was able to remain off antipsychotic medication “without paranoid outbreaks”.

Lastly, McNiel-Gauthier et al. [27] described a 32 year old male who was referred to psychiatric services due to “behavioral outbursts” and “odd beliefs”. In adulthood, he developed “ideas of reference” (e.g., in relation to the television) and “non-systematized persecutory delusions” that involved concerns about being “mocked” by others and surveyed from parked cars. Relatedly, he experienced visual and auditory illusions but no frank hallucinations. Such symptoms were described as occurring daily and episodically, and were reported to be worse after watching television and using the computer. He was also oppositional and aggressive in this context. Lorazepam was initially “mildly successful” in helping “control these outbursts”, but led to sedation. Aripiprazole 2 mg/day was trialed and after three weeks he exhibited a “moderately reduced frequency of ideas of reference” and became “less preoccupied” with his persecutory delusions. While his global improvement score was “2” (“much improved”), he still rarely exhibited “impulsive and disinhibited behaviors”. With respect to EPS, only his dysarthria seemingly worsened. His psychotic symptoms and EPS remained unchanged after three additional months of being on aripiprazole 2 mg/day. While he was weaker, this was attributed to the natural course of the disease and not to the use of aripiprazole. After one year of treatment he experienced a recurrence of “behavioral outbursts” three to four times/week, which was also thought to be related to disease progression. As a result, his dose of aripiprazole was increased to 3 mg/day with a “good response”. Approximately two years after the initial assessment, despite continued neurological deterioration his psychotic and behavioural symptoms remained “stable”. Although periodic “behavioural outbursts” persisted, albeit less frequently, his psychotic symptoms in particular were “much improved” compared to the first assessment. Overall, the authors concluded that aripiprazole therapy resulted in “few residual psychotic symptoms” “without definite medication related motor side effects”.

## Discussion

Interestingly, various antipsychotic medications known to cause EPS were used in most previous case reports, despite KRS being a form of early-onset parkinsonism. In some cases, even typical or first generation antipsychotics, which are particularly problematic in this respect, were used. Although McNiel-Gauthier et al. [27] suggested that low dose aripiprazole was effective without definitive evidence of medication-related EPS in their patient, this was presumably speculative given his progressive motor dysfunction.

Quetiapine and clozapine, which are commonly used in PD when antipsychotic therapy is deemed necessary, were only used in a few cases. Specifically, quetiapine was only used in three patients [21, 24, 25], and its effectiveness and/or impact on motor functioning were only vaguely described by Di Fonzo et al. [21] and Pietrzak et al. [25]. Moreover, the concurrent use of levodopa in both cases [21, 25], as well as the use of olanzapine in one [25], further confounds interpretation of quetiapine’s effectiveness and tolerability in these patients. Clozapine (at a very low dose) has only been used in one published case to date [24]. Although this patient’s hallucinations reportedly improved, it is not clear that their symptoms fully remitted.

Here, we describe a KRS patient carrying a novel homozygous variant in ATP13A2, which was demonstrated to cause a complete loss of protein activity, in accordance with previously characterized ATP13A2 KRS-causing variants [9–13, 3, 5]. Importantly, this is the first report of a patient with KRS whose psychotic symptoms remitted with quetiapine monotherapy. Notably however, Di Fonzo et al. [21] and Pietrzak et al. [25] both utilized a total daily dose of 50 mg, whereas we ultimately had to target a higher dose (300 mg total daily dose). Although quetiapine has so far been well tolerated in our patient, we cannot be certain that it has not contributed to his ongoing motor symptoms, as olanzapine and quetiapine were cross-titrated. That is, at no point was he off of antipsychotic medication entirely; however, he and his family observed significant improvement with respect to his motor functioning following the switch to quetiapine. It should also be noted that at the time of publication, our patient was not yet on dopaminergic therapy of any kind, and it is possible that quetiapine may prove less effective should concurrent levodopa treatment eventually be required. Similarly, quetiapine may prove to be less effective and/or well tolerated as his illness progresses.

Although controlled trials in neurodegenerative parkinsonian disorders have been disappointing [28], quetiapine is nonetheless commonly prescribed for PD psychosis [29] and similarly represents a reasonable option in patients with KRS, given its side effect profile and ease of use. Although clozapine has more evidence in the treatment of PD psychosis [30], we did not pursue a clozapine trial given that our patient’s psychotic symptoms remitted with quetiapine therapy. Although clozapine can also be considered, particularly when psychotic symptoms are treatment resistant and provided no medical contraindications exist, its use is limited by the possibility of rare but potentially serious side effects and the related need for regular blood monitoring. It is also worth noting that no previous articles have described the use of pimavanserin in KRS-associated psychosis, despite pimavanserin being the only FDA approved medication for the treatment of PD psychosis in the United States [30]. This was not an option for our patient, as it is currently not available in our region.

## Conclusion

This report characterizes a homozygous novel loss-of-function ATP13A2 variant in an individual with KRS. This is also the first description of psychotic symptoms fully remitting in response to quetiapine monotherapy in KRS. Given its lower propensity to cause EPS, quetiapine should be considered in the management of KRS-associated psychosis when antipsychotic therapy is deemed necessary.

## Data Availability

All datasets generated or analyzed in this study can be found through the Zenodo depository. All experimental protocols can be found on protocols.io.

## Acknowledgements

None

## Ethics Statement

The patient’s legal guardian has provided written informed consent for the publication of this report, and the patient himself has assented. The University of Calgary Conjoint Health Research Ethics Board (CHREB) has confirmed that this study does not require REB approval.

## Author Contributions

Mark Colijn (psychiatrist), Greg Montgomery (psychiatrist), Justyna Sarna (neurologist), and Billie Au (clinical geneticist) were all involved in the assessment and treatment of the patient and contributed to the development and writing of the manuscript. Peter Vangheluwe, Stephanie Vrijsen, Rania Abou El Asrar, Marine Houdou, and Chris Van den haute were all involved in the design and completion of the biochemical analysis of the genetic variant and also collectively contributed to the writing of the corresponding portions of the manuscript.

## References

1. Gregory A, Hayflick S. Neurodegeneration with Brain Iron Accumulation Disorders Overview. In: Adam MP, Mirzaa GM, Pagon RA, Wallace SE, Bean LJH, Gripp KW, et al., editors. GeneReviews((R)). Seattle (WA): 1993.

2. Yang X, Xu Y. Mutations in the ATP13A2 gene and Parkinsonism: a preliminary review. Biomed Res Int. 2014;2014:371256.

3. Estrada-Cuzcano A, Martin S, Chamova T, et al. Loss-of-function mutations in the ATP13A2/PARK9 gene cause complicated hereditary spastic paraplegia (SPG78). Brain. 2017 Feb;140(2):287–305.

4. Azfar M, van Veen S, Houdou M, et al. P5B-ATPases in the mammalian polyamine transport system and their role in disease. Biochim Biophys Acta Mol Cell Res. 2022 Dec;1869(12):119354.

5. van Veen S, Martin S, Van den Haute C, et al. ATP13A2 deficiency disrupts lysosomal polyamine export. Nature. 2020 Feb;578(7795):419-24.

6. Vrijsen S, Houdou M, Cascalho A, et al. Polyamines in Parkinson’s Disease: Balancing Between Neurotoxicity and Neuroprotection. Annu Rev Biochem. 2023 Jun 20;92:435–64.

7. Fiori LM, Turecki G. Implication of the polyamine system in mental disorders. J Psychiatry Neurosci. 2008 Mar;33(2):102–10.

8. Vrijsen S, Besora-Casals L, van Veen S, et al. ATP13A2-mediated endo-lysosomal polyamine export counters mitochondrial oxidative stress. Proc Natl Acad Sci U S A. 2020 Dec 8;117(49):31198–207.

9. Ning YP, Kanai K, Tomiyama H, et al. PARK9-linked parkinsonism in eastern Asia: mutation detection in ATP13A2 and clinical phenotype. Neurology. 2008 Apr 15;70(16 Pt 2):1491-3.

10. Dehay B, Ramirez A, Martinez-Vicente M, et al. Loss of P-type ATPase ATP13A2/PARK9 function induces general lysosomal deficiency and leads to Parkinson disease neurodegeneration. Proc Natl Acad Sci U S A. 2012 Jun 12;109(24):9611–6.

11. Grunewald A, Arns B, Seibler P, et al. ATP13A2 mutations impair mitochondrial function in fibroblasts from patients with Kufor-Rakeb syndrome. Neurobiol Aging. 2012 Aug;33(8):1843 e1-7.

12. Podhajska A, Musso A, Trancikova A, et al. Common pathogenic effects of missense mutations in the P-type ATPase ATP13A2 (PARK9) associated with early-onset parkinsonism. PLoS One. 2012;7(6):e39942.

13. Usenovic M, Tresse E, Mazzulli JR, et al. Deficiency of ATP13A2 leads to lysosomal dysfunction, alpha-synuclein accumulation, and neurotoxicity. J Neurosci. 2012 Mar 21;32(12):4240–6.

14. Martin S, van Veen S, Holemans T, et al. Protection against Mitochondrial and Metal Toxicity Depends on Functional Lipid Binding Sites in ATP13A2. Parkinsons Dis. 2016;2016:9531917.

15. Houdou M, Jacobs N, Coene J, et al. Novel Green Fluorescent Polyamines to Analyze ATP13A2 and ATP13A3 Activity in the Mammalian Polyamine Transport System. Biomolecules. 2023 Feb 9;13(2).

16. Williams DR, Hadeed A, al-Din AS, et al. Kufor Rakeb disease: autosomal recessive, levodopa-responsive parkinsonism with pyramidal degeneration, supranuclear gaze palsy, and dementia. Mov Disord. 2005 Oct;20(10):1264–71.

17. Schneider SA, Paisan-Ruiz C, Quinn NP, et al. ATP13A2 mutations (PARK9) cause neurodegeneration with brain iron accumulation. Mov Disord. 2010 Jun 15;25(8):979–84.

18. Estiar MA, Leveille E, Spiegelman D, et al. Clinical and genetic analysis of ATP13A2 in hereditary spastic paraplegia expands the phenotype. Mol Genet Genomic Med. 2020 Mar;8(3):e1052.

19. Odake Y, Koh K, Takiyama Y, et al. Identification of a novel mutation in ATP13A2 associated with a complicated form of hereditary spastic paraplegia. Neurol Genet. 2020 Oct;6(5):e514.

20. Satolli S, Di Fonzo A, Zanobio M, et al. Kufor Rakeb syndrome without gaze palsy and pyramidal signs due to novel ATP13A2 mutations. Neurol Sci. 2023 Oct;44(10):3723–25.

21. Di Fonzo A, Chien HF, Socal M, et al. ATP13A2 missense mutations in juvenile parkinsonism and young onset Parkinson disease. Neurology. 2007 May 8;68(19):1557–62.

22. Chien HF, Rodriguez RD, Bonifati V, et al. Neuropathologic Findings in a Patient With Juvenile-Onset Levodopa-Responsive Parkinsonism Due to ATP13A2 Mutation. Neurology. 2021 Oct 19;97(16):763–66.

23. Behrens MI, Bruggemann N, Chana P, et al. Clinical spectrum of Kufor-Rakeb syndrome in the Chilean kindred with ATP13A2 mutations. Mov Disord. 2010 Sep 15;25(12):1929–37.

24. Abbas MM, Govindappa ST, Sheerin UM, et al. Exome Sequencing Identifies a Novel Homozygous Missense ATP13A2 Mutation. Mov Disord Clin Pract. 2017 Jan-Feb;4(1):132-35.

25. Pietrzak A, Badura-Stronka M, Kangas-Kontio T, et al. Clinical and ultrastructural findings in an ataxic variant of Kufor-Rakeb syndrome. Folia Neuropathol. 2019;57(3):285–94.

26. Balint B, Damasio J, Magrinelli F, et al. Psychiatric Manifestations of ATP13A2 Mutations. Mov Disord Clin Pract. 2020 Oct;7(7):838–41.

27. McNeil-Gauthier AL, Brais B, Rouleau G, et al. Successful treatment of psychosis in a patient with Kufor-Rakeb syndrome with low dose aripiprazole: a case report. Neurocase. 2019 Jun-Aug;25(3-4):133-37.

28. Desmarais P, Massoud F, Filion J, et al. Quetiapine for Psychosis in Parkinson Disease and Neurodegenerative Parkinsonian Disorders: A Systematic Review. J Geriatr Psychiatry Neurol. 2016 Jul;29(4):227–36.

29. Friedman JH. Pharmacological interventions for psychosis in Parkinson’s disease patients. Expert Opin Pharmacother. 2018 Apr;19(5):499–505.

30. Rissardo JP, Durante I, Sharon I, et al. Pimavanserin and Parkinson’s Disease Psychosis: A Narrative Review. Brain Sci. 2022 Sep 23;12(10).

